# Assessing the diagnostic impact of blood transcriptome profiling in a pediatric cohort previously assessed by genome sequencing

**DOI:** 10.1101/2024.12.03.24317221

**Authors:** Huayun Hou, Kyoko E. Yuki, Gregory Costain, Anna Szuto, Sierra Barnes, Arun K. Ramani, Alper Celik, Michael Braga, Meagan Gloven-Brown, Dimitri J. Stavropoulos, Sarah Bowdin, Ronald D Cohn, Roberto Mendoza-Londono, Stephen W. Scherer, Michael Brudno, Christian R. Marshall, M. Stephen Meyn, Adam Shlien, James J. Dowling, Michael D. Wilson, Lianna Kyriakopoulou

## Abstract

Despite advances in diagnostic testing and genome sequencing, the majority of individuals with rare genetic disorders remain undiagnosed. As a complement to genome sequencing, transcriptional profiling can provide insight into the functional consequences of DNA variants on RNA transcript expression and structure. Here we assessed the utility of blood derived RNA-seq in a well-studied, but still mostly undiagnosed, cohort of individuals who enrolled in the SickKids Genome Clinic study. This cohort was established to benchmark the ability of genome sequencing technologies to diagnose genetic diseases and has been subjected to multiple analyses. We used RNA-seq to profile whole blood RNA expression from all probands for whom a blood sample was available (n=134). Our RNA-centric analysis included differential gene expression, alternative splicing, and allele specific expression. In one third of the diagnosed individuals (20/61), RNA-seq provided additional evidence supporting the pathogenicity of the variant found by prior DNA-based analyses. In 2/61 cases, RNA-seq changed the GS-derived genetic diagnosis (*EPG5* to *LZTR1* in an individual with a Noonan syndrome-like disorder) and discovered an additional relevant gene (*CEP120* in addition to *SON* in an individual with ZTTK syndrome). In ∼7% (5/73) of the undiagnosed participants, RNA-seq provided at least one plausible, potentially diagnostic candidate gene. This study illustrates the benefits and limitations of using whole-blood RNA profiling to support existing molecular diagnoses and reveal candidate molecular mechanisms underlying undiagnosed genetic disease.

## INTRODUCTION

Exome and genome sequencing (GS) technologies have transformed our ability to identify causal genetic variants and make diagnoses for rare diseases (e.g. (1); reviewed by (2,3)). Despite this progress, more than half of individuals with a genetic disorder remain without a diagnosis. This can be partly attributed to our limited understanding of the downstream consequences of genetic variation and the lack of functional evidence for its interpretation. RNA sequencing (RNA-seq) has the potential to identify clinically relevant aberrations and provide a high throughput functional platform for those variants related to RNA expression levels and/or RNA processing.

Several studies have used transcriptome profiling to assess its potential diagnostic utility in rare disorders (4–15). For example, RNA-seq on muscle biopsies identified molecular diagnoses in up to 35% of individuals with neuromuscular disorders (8,9,12). RNA-seq using fibroblasts yielded a diagnostic rate of 10-16% in individuals with mitochondrial diseases (13,14). Fresard et al. (2019) demonstrated that RNA-seq in whole blood increased the absolute diagnostic rate by 7.5% in a phenotypically heterogeneous cohort with suspected rare diseases (11). While these studies support the ability of RNA-seq to improve diagnostic rates, there are few studies benchmarking the RNA-seq yield when performed as an adjunct to genome sequencing (10).

In this study, we assessed the utility of RNA-seq for decreasing uncertainty and increasing the yield of GS-based diagnoses. To do this we performed RNA-seq on RNA isolated from peripheral blood samples donated by children and youth with suspected genetic disorders who enrolled in the Genome Clinic at the Hospital for Sick Children (Toronto, ON, Canada).

This longitudinal study was designed to test the diagnostic and predictive use of GS (16,17). The participants presented with a spectrum of complex clinical phenotypes such as epilepsy, global developmental delay and multiple congenital anomalies and had previously been tested by standard of care genetic testing. After genome sequencing and subsequent reanalyses, the diagnostic rate was 41% (16–18). Given the clinical work up and intense focus on genome analysis in this cohort, we reasoned that performing RNA-seq on all individuals who provided blood samples would be an important and unique opportunity to examine the concordance between genome-based diagnoses and RNA-seq as well as ask whether new potential candidate disease genes related to the individual’s phenotype could be revealed by RNA-seq. In this study we focused on two clinically meaningful scenarios, where we asked if transcriptomic data could: 1) provide functional evidence for several classes of suspected/known diagnostic genetic variants identified by GS; and 2) identify plausible candidate genes for individuals for whom GS was initially non-diagnostic.

## MATERIALS AND METHODS

### Cohort description

The cohort in this study was derived from the SickKids Genome Clinic which consisted of two studies that collected samples over a 2-year period (April 2013 to June 2015) (16,17,19). The participants presented with a spectrum of complex clinical indications including epilepsy, global developmental delay, and multiple congenital anomalies, which represent the most common indications for genetic testing. The SickKids Genomic Clinic participants first underwent genetic testing that met the standard of care provided at the time and were subsequently examined using genome sequencing. The first study cohort (which compared the diagnostic rate of genome sequencing to traditional chromosomal microarray) involved 100 children with suspected genetic disorders (16). The second cohort (which compared genome sequencing to targeted gene panels) involved 103 children (17). This study was approved by the Research Ethics Board at The Hospital for Sick Children and informed consent was obtained for all participants (REB number 1000037726). Case IDs are de-identified and are not known to anyone outside the research group.

Individual’s phenotypic data was used in this study, to prioritize splicing and differential expression events, were previously captured in PhenoTips (http://www.phenotips.com) (20).

Phenotypic information is represented using the Human Phenotype Ontology (HPO) (21). Participant data also included data regarding molecular genetic testing, including single gene or gene panels, extracted from the electronic medical records. Chemistry tests (blood and urine), enzymatic studies, muscle biopsies, and medical imaging, when available, were also included as previously described by both Stavropoulos et al. (2016) and Lionel et al. (2018) (16,17).

### RNA isolation, library preparation, and sequencing

RNA from the 134 probands were extracted from blood collected in PAXGene tubes using an automated QIAsymphony PAXGene blood RNA kit (Qiagen). RNA was quantified using the Qubit Fluorometer (Thermo Fisher Scientific) High Sensitivity Assay, and sample purity was checked using the Nanodrop (Thermo Fisher Scientific) OD 260/280 ratio. Following the manufacturer’s recommended protocol, 250 ng of total RNA spiked with the Spike-in RNA variants of the isoform (SIRV) Set 3 (Lexogen), such that spike-ins were ∼1% of the mRNA, were used as input material for library preparation. SIRV set3 includes 92 ERCCs and 69 SIRV isoforms corresponding to seven SIRV genes, allowing the assessment of an RNA-seq platform’s ability to detect a range of RNA concentrations as well as isoform complexity. Libraries were prepared using an automated NEBNext Ultra II Directional Library Prep Kit for Illumina with polyA isolation at Genome Diagnostics (Department of Laboratory Medicine, The Hospital for Sick Children). Libraries were assessed using the Bioanalyzer DNA High Sensitivity chips (Agilent Technologies, Santa Clara, CA) and quantified by quantitative polymerase chain reaction using the Kapa Library Quantification Illumina/ABI Prism Kit protocol (KAPA Biosystems, Roche, Basel, Switzerland). Libraries were pooled in equimolar quantities and paired-end sequenced on an Illumina NovaSeq6000 system, following Illumina’s recommended protocol, to generate paired-end reads of 150 bases in length at The Centre for Applied Genomics, The Hospital for Sick Children, Toronto, Canada. A median of 116 million paired-end reads per library was obtained.

### RNA-seq data analysis and annotation

A customized RNA-seq processing pipeline was used for read alignment, quality control (QC), identification of expression and splicing aberrations, and variant calling. Raw sequencing reads were aligned using STAR (22) (v2.6.1c) to the combined genome of hg19 (1000 genomes reference genome, hs37d5) and the spike-in sequences (SIRVome, SIRV set3). Gene annotation was obtained from Ensembl (v75) and was combined with SIRVome transcript annotation obtained from the manufacturer (https://www.lexogen.com/sirvs/download/). Additional gene-disease association annotations were obtained from various sources: OMIM gene accession IDs and disease descriptions (Ensembl v75); HPO gene-disease association ( (https://hpo.jax.org/data/annotations); numbers of ClinVar variants for each gene (https://ftp.ncbi.nlm.nih.gov/pub/clinvar/tab_delimited/gene_specific_summary.txt); and Orphanet (http://www.orphadata.org/) in November 2019. Fastqc (23) (v0.11.5), RNA-seQC (24) (v2.3.5), Picard (https://broadinstitute.github.io/picard/) (v2.18.0) Markduplicates and CollectRnaSeqMetrics, supplemented with customized scripts were used to collect various QC metrics. Gene and transcript expression level quantification was performed using RSEM (25) (v1.2.22).

### Gene expression outlier analysis

Gene expression outliers were identified by comparing each sample to the rest of the samples in the cohort using R package OUTRIDER (26) (v 1.8.0). First, to minimize the effect of variable hemoglobin content, we removed nine hemoglobin genes (H*BB, HBD, HBG1, HBG2, HBZ, HBM, HBA2, HBA1, HBQ1*). Next, gene read counts estimated by RSEM were filtered for lowly expressed genes (only genes with a 95^th^ percentile RPKM >1 were used). OUTRIDER results for samples 230373, 230577 and 239099 were not analyzed as they were identified outliers after PCA analysis using OUTRIDER normalized counts (Supplementary Figure 1). Subsequent examination showed these samples have suboptimal library quality. For each sample, genes with either an adjusted p value < 0.05 or an absolute z-score ≥ 3 were reported as expression outliers and further examined manually.

### Splicing outlier analysis

To identify aberrant splicing events, we adopted the approach described by Fresard et al. (11). Briefly, we used junction quantification by STAR (*SJ.out file). Only junctions with more than 5 uniquely mapped reads, were considered in the analysis. Next, we calculated a junction coverage score, defined as the number of reads mapping to a junction of interest divided by the total number of reads mapping to other junctions that share a splicing donor or acceptor site with the junction of interest. We then calculated the z-score of junction coverage score for each junction within the study cohort. Junctions with an absolute z-score ≥ 2 were further examined.

In addition, we also applied the aberrant splicing module from the Detection of RNA Outlier Pipeline (DROP) (27) with default settings as a complementary approach to identify aberrant splicing events. Specifically, the DROP pipeline uses R package FRASER (28) which assesses the statistical significance of aberrant splicing events in addition to reporting z-scores, providing additional criteria for prioritization. Splicing events were visualized using ggsashimi (29). Gene models were visualized using R package Gviz based on Ensembl (v75) gene annotation (30).

### Allele specific expression

Allele-specific expression (ASE) events was detected using a method adapted from the “MAE” module of the DROP pipeline. Briefly, for each sample, heterozygous SNPs identified from GS were first filtered for quality as previously described (16,17) and further filtered to retain variants that are only identified in one sample in our internal batch. “ASEReadCount” from GATK (31) (v4.0.1.2) was used to count the number of RNA-seq reads mapping to reference or alternative alleles of filtered SNPs. DESeq2 (32) (v1.44) was then used to test for significant bias towards reference or alternative alleles. ASE events with an alternative allele read ratio > 0.8 and adjusted p values < 0.05 were further examined.

### Interpretation workflow

Splicing junctions were prioritized first using technical filters, including z-scores, number of junction reads, junction coverage ratio, etc., followed by the biological significance of the genes containing the candidate junctions. Specifically, genes with HPO terms corresponding to each participant’s disease presentation were prioritized, followed by genes with any disease associations, and genes with no known disease associations. All splicing junctions that were considered clinically significant were visually inspected using the Integrated Genomics Viewer (IGV) (33). Expression outliers and ASE genes were prioritized using z-scores and the adjusted p value for each sample. Similarly, genes with relevant HPO terms or disease associations were prioritized.

## RESULTS

The cohort for this study consisted of all individuals from the SickKids Genome Clinic for whom a primary blood sample was available (84/100 from Stavropoulos et al. 2016 (16) and 50/103 from Lionel et al. 2018 (17)). This provided us with a cohort of 134 participants (75 males and 59 females) between 0 and 18 years of age at the time of sample collection (median 6 years; Supplementary Table 1). These individuals displayed a wide array of symptoms described by 453 unique HPO terms across the cohort. The most observed phenotypes were epilepsy (29%), global developmental delay (25%), multiple congenital abnormalities (19%), connective tissue (5%), eye (6%), behaviour/cognition - other (3%), neurological - other (2%), immune (2%), cardiovascular (1%) and metabolic including mitochondrial (9%; Table 1 and Supplementary Table 2).

**Table 1:**
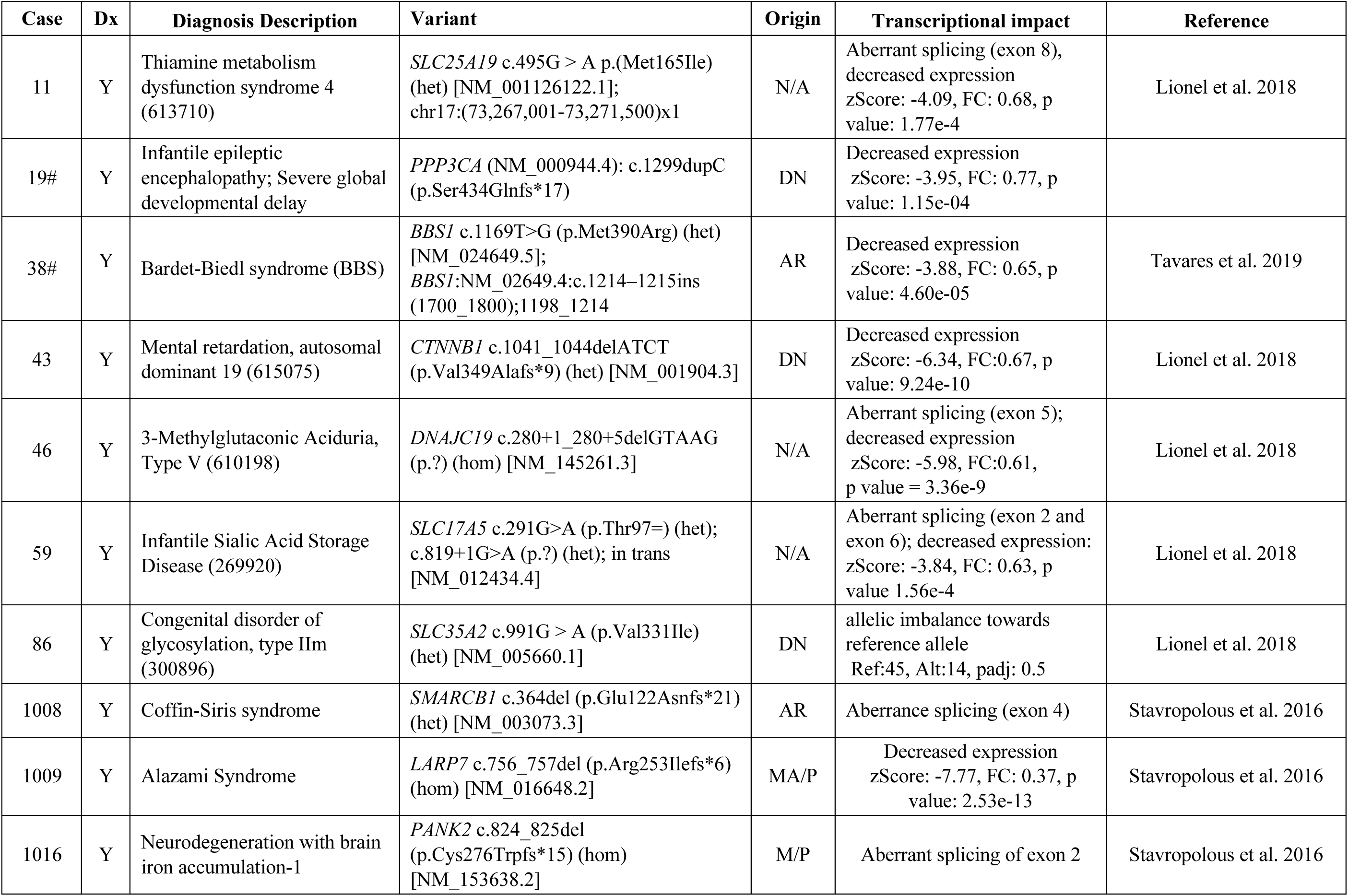
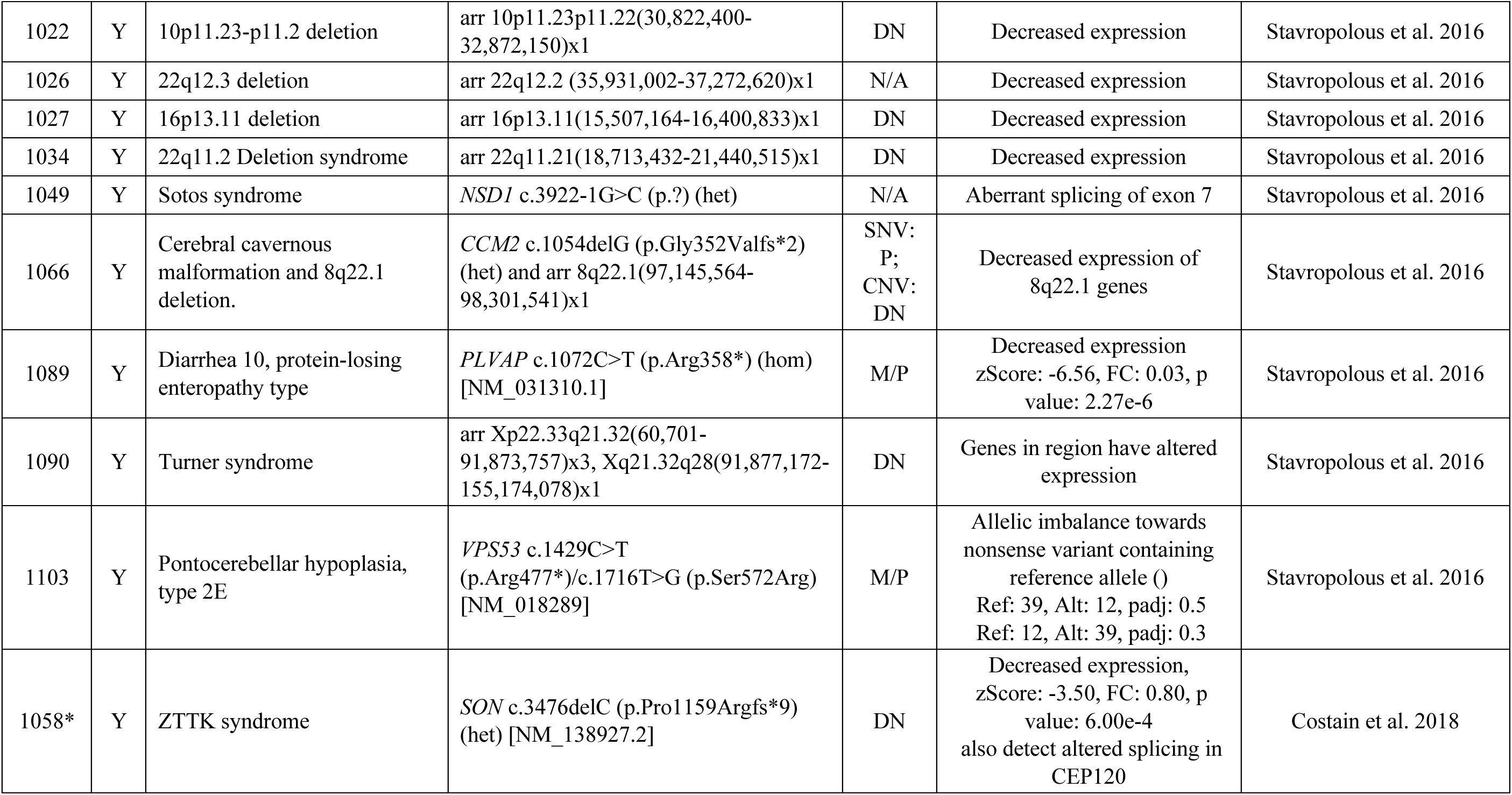
Transcriptional impact of disease genes detected in blood. The case ID, presence of a diagnosis (Dx). Cases included in cohort from Lionel et al. 2018 are designated by #; Cases included in the cohort from Stavropolous et al. 2016 are designated with *; het: heterozygous; hom: homozygous; Origin of mutation abbreviations: M: maternal; P: paternal; DN: *de novo*; N/A: not available.

Based on prior published analyses we split this cohort into two groups: Group 1 consisted of 61 individuals (35 male and 26 female) who received a diagnostic DNA variant(s) in the initial GS based studies (16–18). Group 2 consisted of the remaining 73 individuals (40 male and 33 female), who have yet to receive a molecular genetic diagnosis. The logic behind partitioning this well studied cohort into two groups was that it would allow us to: a) assess the relevance of RNA-seq alone in identifying/corroborating existing diagnoses (Group 1); and b) suggest genes and possible pathogenic variants related to the primary indication for referral in the unsolved cases (Group 2).

### Developing and applying a scalable and reproducible RNA-seq library preparation method to 134 whole blood samples

Compared to genome sequencing, considerable technical variability can occur when performing RNA-seq. Our long-term objective is to establish a clinically validated RNA-seq procedure that can run in our clinical lab so that RNA-seq samples prepared over extended periods of time will show minimal batch effects. We established a semi-automated RNA-seq library preparation protocol with a workflow suitable for clinical laboratory testing. The sample preparation protocol included synthetic spike-in controls that allow us to assess the quality of the library preparation (ERCC + SIRV controls; see Methods and Figure 1A). We chose not to pursue globin depletion strategies with the rationale that this would increase the variability in the library preparation step. Instead, we implemented deeper sequencing of libraries to ensure sufficient gene detection in samples where globin mRNA is high.

**Figure 1.**
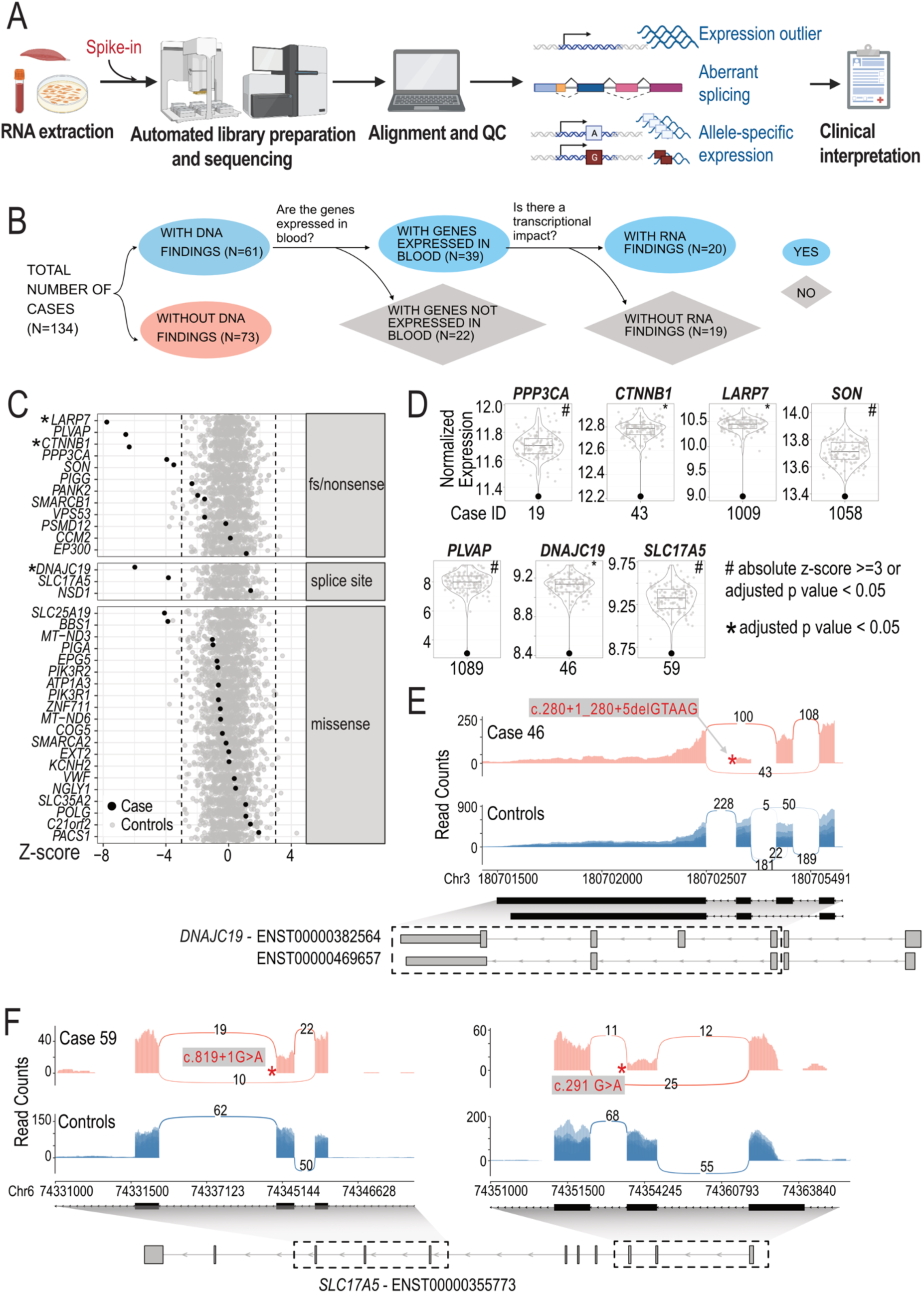
Effects of pathogenic or likely pathogenic SNVs and indels on gene expression. A. RNA-seq-based clinical diagnosis workflow Created in https://BioRender.com. B. Flowchart showing the breakdown of samples. C. Detection of candidate genes as expression outliers. Each row represents one candidate gene. Each dot represents the z-scores of the corresponding gene in all samples. Black: cases in which the corresponding gene was a candidate; Grey: rest of the cohort as control samples. Asterisks indicate that the candidate gene is also detected as significant (adjusted p value < 0.05). Dashed lines indicated z-score of –3 or 3. Genes are separated into different panels based on the type of mutations they harbor. D. Violin and boxplot showing normalized gene expression of selected candidate genes across the cohort. Y axis: log2 normalized read counts of the target gene in all samples in the cohort. Each dot represents one sample. Highlighted dot represents the case of interest. Case IDs are shown below each plot. Number sign: the gene is considered an outlier in the analysis (absolute z-score >=3 or adjusted p value < 0.05); asterisk: the gene is detected as significant outlier (adjusted p value < 0.05). Impact of pathogenic variants in *DNAJC19* (E) and *SLC17A5* (F) on splicing and gene expression. Sashimi plots show reads and junctions in the affected region for case (red track) and control samples (blue track, n = 10, randomly selected from the cohort) with the DNA variant labeled with an asterisk. Structures of relevant transcripts are shown with UTRs shown as thinner boxes. Exons shown in the sashimi plots are highlighted with a dashed box.

### Using RNA-seq to reveal the impact of variants detected in disease genes obtained through prior genomic testing

Despite blood samples having been archived for 3-5 years, we obtained RNA libraries suitable for subsequent sequencing for all samples. The resulting RNA-seq libraries were sequenced to a median of 116 million reads with a median of 78% of the reads mapped uniquely to the hg19 genome. We detected a median of 12,071 genes (TPM >1) and 176,284 junctions (>=5 reads supporting a junction) per sample across our cohort. A full list of quality control metrics for each sample can be found in Supplementary Table 1.

We focused on the 61 individuals for whom genomic variants in the initial studies and subsequent re-analysis had been considered as receiving a diagnosis (16–18) (Group 1). These 61 cases were comprised of 52 cases with SNVs or indels, six cases with structural variants (SVs)/copy number variants (CNVs), and three cases with both candidate SNVs/indels and SV/CNVs. In 39 of the 61 cases, the disease-causing genes underlying the diagnosis were detectable in our blood RNA-seq data. RNA-seq revealed that in 20/39 cases the variants influenced transcription (altered expression, splicing, or allele-specific expression; Table 1, Figure 1B).

### RNA expression changes in previously identified disease genes

We examined the effect of SNVs or indels on gene expression for genes that we detected by whole blood RNA-seq. This variation included missense variants, splice site variants, frameshift or nonsense variants (Figure 1C, Table 1, Supplementary Table 2). We found decreased gene expression levels in genes from 9 cases, including 5 cases with nonsense or frameshift variants associated with autosomal dominant (AD) Infantile Epileptic Encephalopathy with Severe Global Developmental delay (*PPP3CA*), autosomal recessive (AR) Alazami syndrome (*LARP7*), AD Mental Retardation Autosomal Dominant 19 (*CTNNB1*) and AR Diarrhea 10, protein-losing enteropathy type (*PLVAP*), AD ZTTK syndrome (*SON*) (Figure 1C-F, Table 1).

RNA-seq also gave us an opportunity to assess the gene expression consequences of missense variants. While it is typically assumed that missense variants would not overtly influence gene expression levels, genome sequencing alone cannot determine this. We found that 19/21 of the diagnosed cases with a missense variant showed no obvious effect on gene expression. The genes from the two participants with a diagnostic missense variant showed decreased gene expression (z-score < -3) were *SLC25A19* (Case 11) and *BBS1* (Case 38) (Figure 1C, Table 1). The *SLC25A19* result can be explained by the fact that in addition to a missense variant, the individual also possessed a heterozygous deletion that removes part of the last intron and the last exon of *SLC25A19* (Table 1). Case 38 did not have an initial explanation for the observed reduced *BBS1* expression and the clinical data we used (16–18) only reported the heterozygous pathogenic missense variant (NM_024649.5) c.1169T>G (p.Met390Arg).

However, additional consultation revealed that subsequent genome reanalysis study at SickKids independently determined that a retrotransposon insertion occurred *in trans* to the heterozygous pathogenic missense variant resulting in a premature termination codon (34). Our RNA-seq analysis pipeline does not readily allow for the automated detection of such insertions and deletions. We did not initially flag these reads for further inspection due to the low expression of *BBS1* in blood. Manual inspection of the RNA-seq reads identified reads mapping to the exon and the retrotransposon sequence.

### Evidence for RNA splicing changes in previously identified disease genes

We observed splicing related consequences in five individuals:

***Case 46*:** aberrant splicing was present in an individual with a suspected mitochondrial disorder carrying a homozygous deletion occurring in the splice donor site of exon 5 of the *DNAJC19* gene (NM_145261.3) c.280+1_280+5delGTAAG which is associated with 3- Methylglutaconic Aciduria Type V. Interestingly, this variant results in two novel junctions, one causing the skipping of exon 5 and the other causing the skipping of both exon 4 and 5 (Figure 1E). Both events are predicted to result in a frameshift. In addition, reduced expression of *DNAJC19* gene was also observed (Figure 1D). While this variant was predicted to affect a canonical splice site, RNA-seq demonstrates a functional consequence of this deletion *in vivo*.

***Case 59:*** aberrant splicing was confirmed in a case of Infantile Sialic Acid Storage disease where two rare variants in trans (confirmed by parental studies) were identified in the *SLC17A5* gene (NM_012434.4) c.819+1G>A; c.291G>A p.(Thr97=). Both variants were predicted to influence splicing by *in silico* predictions. RNA-seq analyses also confirm that aberrant splicing occurs and showed skipping of exon 2 (c.291G>A p.Thr97=) and exon 6 (c.819+1G>A p.?) (Figure 1F). Both of these events are predicted to result in a frameshift.

Moreover, these aberrant splicing events were associated with reduced levels of the *SLC17A5* transcripts indicating the occurrence of nonsense mediated decay (NMD) (Table 1; Figure 1D). ***Case 1008*:** a rare heterozygous variant in the *SMARCB1* gene (NM_003073.3) c.364del (p.Glu122Asnfs*21) was previously identified in an individual clinically diagnosed with Coffin-Siris syndrome. This variant was predicted to cause a frameshift in this haploinsufficient gene. Our RNA-seq analysis did not reveal a significantly reduced the expression of this gene that would be expected from NMD (z-score: -1.53, fold change: 0.89, p value: 0.12). However, RNA splicing analysis showed a decreased usage of exon 4 via exon skipping (Supplementary Figure 2A). Importantly c.364del (p.Glu122Asnfs*21) occurs adjacent to the splice acceptor site, two base pairs from the start of exon 4. Skipping of exon 4 results in an in-frame truncation of the SMARCB1 protein, the functional consequences of which remain unknown.

***Case 1016:*** a homozygous frameshift deletion was identified in exon 2 of the *PANK2* gene (NM_153638.2) c.824_825del (p.Cys276Trpfs*15)) in an individual diagnosed with Neurodegeneration with Brain Iron Accumulation-1. Based on our RNA-seq data, three isoforms are estimated to account for > 90% of *PANK2* expression (ENST00000336066, ENST00000497424, and ENST00000610179). RNA-seq estimated an increased usage of ENST00000336066 (65% vs. 52% cohort median) and decreased usage of ENST00000610179 (0 vs. 16% cohort median). Although we only observed a small effect on the overall gene expression level (z-score: -1.97, fold change: 0.88, p value: 0.07; Supplementary Figure 2B), the decreased usage of the transcript expressing the frameshift is likely due to NMD. The shift in usage to an isoform that skips the frameshift may be a compensatory mechanism for maintaining levels of *PANK2* expression. The impact at the protein level remains to be seen.

***Case 1049:*** a heterozygous canonical splice site variant was identified in intron 6 of the *NSD1* gene (NM_022455.4) c.3922-1G>C (p.?) which is associated with Sotos Syndrome. Our RNA-seq analysis detects two aberrant splicing events: The first involves skipping of exon 7 and increased usage of the resulting isoform (ENST00000375350) which is an established frameshift splice isoform annotated as an NMD transcript in the Ensembl database. However, our data shows that the *NSD1* variant increased the fraction of this isoform and that it escaped nonsense mediated decay as the levels of *NSD1* expression where slightly higher in the proband relative to controls. Supporting this observation, a recent genome-wide screen for NMD escaping transcripts also identified ENST000000375350 as a NMD escaping frameshift variant (35). The second splicing event involves the use of an alternative splice acceptor site 5 bp into exon 7 and is predicted to result in a frameshift (Supplementary Figure 2C). The detection of the aberrant transcripts suggests that they may be stable enough to be translated into a truncated protein with little residual or altered function.

### Evidence for allele-biased gene expression in previously assigned disease genes

We observed allele-biased expression in two individuals:

***Case 86:*** A heterozygous missense variant in gene *SLC35A2* (NM_005660.1: c.991G>A (p.Val331Ile)), associated with X-linked dominant Congenital disorder of glycosylation type I, showing allelic imbalance favoring the reference allele (47/61 reads, 74%). This could be due to skewed X-inactivation in different tissues as has been previously reported (36).

***Case 1103:*** Two variants *in trans* in the *VPS53* gene (NM_018289.3) a c.1429C>T (p.Arg477*) and a c.1716 T>G (p.Ser572Arg), associated with Pontocerebellar hypoplasia, type 2E, showed allelic bias towards the p.Arg477 reference residue (42/55 reads; 74%) and the p.Ser572Arg alternative allele (43/58 reads; 74%) respectively. This suggests that NMD of transcript harbouring the nonsense variant is occurring.

### Detecting gene expression outliers at disease associated CNVs

CNVs contribute significantly to rare diseases in human (e.g. (37–40) and that the effect of a CNV on gene expression can be detected in the general population, cancer and in rare diseases (41–48). We examined the nine cases that carried diagnostic or partially diagnostic CNVs and asked if genes overlapping these CNVs show expression changes. In seven cases, these CNVs overlap one or more genes that are sufficiently detected in our cohort. Among these cases, six harbored heterozygous deletions that range from 4 kb to 1.3 Mb, overlapping 3 to 42 genes expressed in blood. In all six cases, we observed decreased expression of genes within the deleted regions as indicated by the decreased average z-score (compared to the rest of the cohort) of their expression (Figure 2A). However, despite the consistent gene dosage change, the expression of genes within the same CNVs were not impacted uniformly (see Supplementary Table 3 for complete lists of outlier genes within CNVs for each sample). One individual with 22q11.2 deletion syndrome harbors the well-characterized heterozygous ∼3MB deletion on chromosome 22 overlapping 137 genes. We focused on the expression of 34 protein-coding genes in this region and found that 24 of them showed decreased expression (z-score <= -3) with fold changes ranging from 0.37 to 0.69 (Figure 2B-C, Supplementary Table 3).

**Figure 2.**
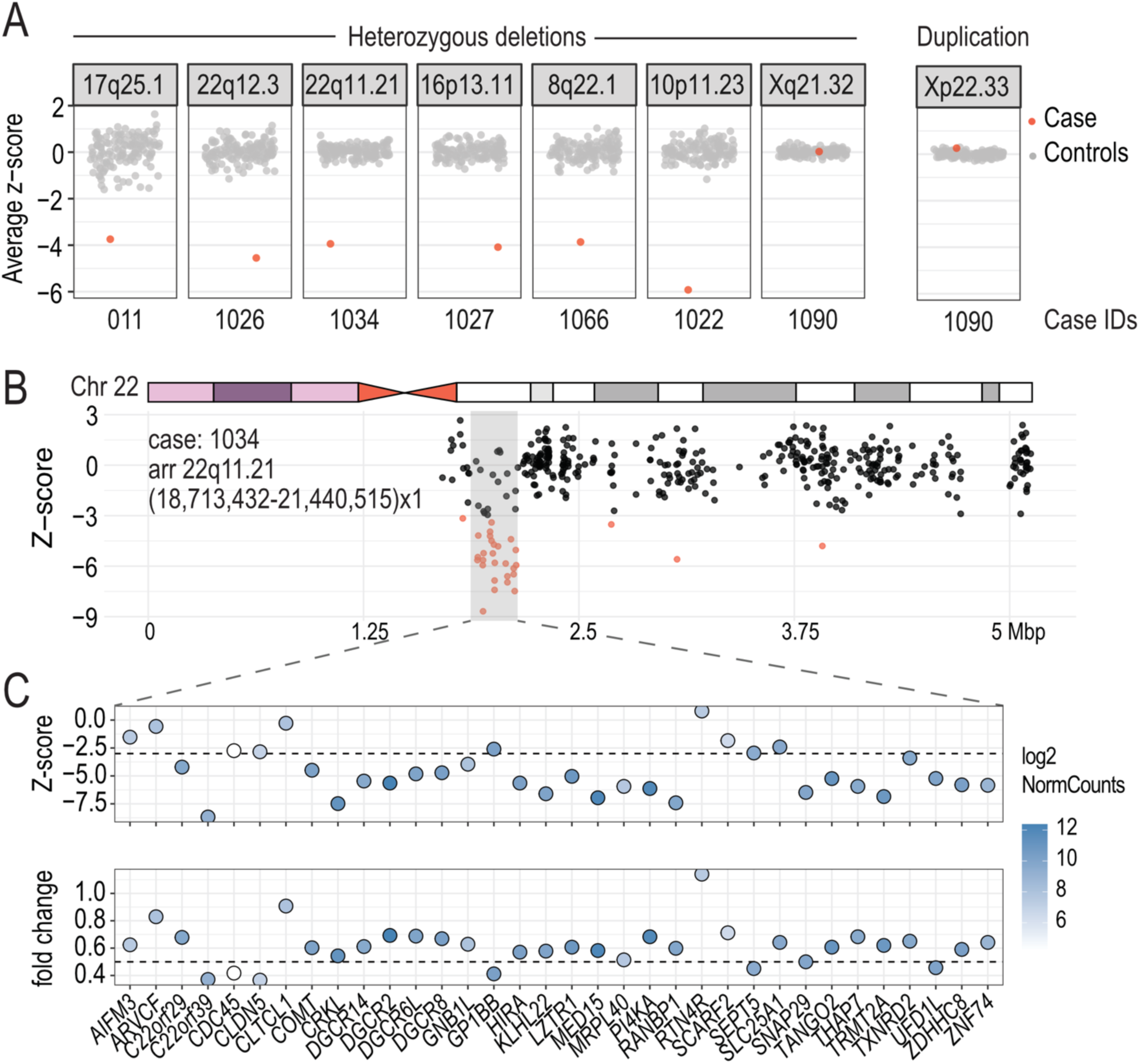
Modulation of gene expression by CNVs. **A.** Z-score summary of genes within pathogenic CNVs. Y axis: average z-score of genes in pathogenic CNVs. Each dot represents one sample. Red: sample in which the pathogenic CNV is detected. Genomic locations of CNVs are shown as panel headers and case IDs are shown below. **B. Z-scores of all genes on chromosome 22 in case 1034.** An **i**deogram of human chromosome 22 is shown. The heterozygous deleted region is highlighted in grey. Gene start is used as the proxy for gene location. Genes with an absolute z-score >= 3 are highlighted in red. **C. Expression changes of genes within the CNV**. Dot plots show z-scores and fold-changes of detected protein-coding genes within the deleted region. Color represents the expression level (log2 normalized counts) of genes in this sample. Genes are ordered by genomic coordinates.

Finally, the last CNV case (Case 1022) is an individual that carried complex X chromosome rearrangements (arr Xp22.33q21.32(60,701-91,873,757)x3, Xq21.32q28(91,877,172-155,174,078)x1) shows congenital anomalies, global developmental delay, severe intellectual disability, seizures, blindness, and deafness (Table 1). Previous studies have shown that despite X chromosome copy number variations, most differentially expressed genes in individuals with Turner Syndrome are on autosomes (reviewed in (49)). To control for X chromosome copy number, we performed gene expression outlier analysis with OUTRIDER using female samples (n = 58) only. We first confirmed that this reanalysis did not change the observation of CNV-overlapping genes in the other cases (Supplementary Figure 3A). Next, we re-examined gene expression changes in this individual. Consistent with previous reports, most genes with expression changes in case 1022 are not X-linked (7 X-linked genes /127 genes with absolute z-score>=3 or adjusted p value < 0.05, Supplementary Table 4). We found that X-linked genes within the duplicated region showed higher expression relative to the rest of the cohort, especially for p-arm genes distal to the centromere (Supplementary Figure 3B-C). In contrast, the expression of genes within the deleted region is comparable to controls (Supplementary Figure 3B-C) which could be explained by skewed X inactivation, which is known to occur with large structural changes on X chromosome (50).

### Identifying splicing and expression outliers in a cohort with no previous molecular findings

There were 73 participants who despite traditional testing and reanalysis remained undiagnosed (16–18). The clinical findings in this cohort included global developmental delay, epilepsy, and multiple congenital anomalies, connective tissue abnormalities, cardiomyopathy, suspected metabolic disorder, and ophthalmological or immune system abnormalities (see Supplementary Table 2). Using the information obtained from our RNA-seq analyses and discussions with clinicians, we identified plausible candidate genes related to the phenotype in five cases (Table 2):

**Table 2:** Candidate genes and additional variants detected in 7 cases.

| Case | Primary indication for referral | RNA-seq Findings | DNA variant | Cohort |
| --- | --- | --- | --- | --- |
| 18 | Global developmental delay and epileptic encephalopathy | *UGDH* decreased expression<br>zScore: -4.15, FC: 0.62, p value: 7.50e-5 | Not detected | Lionel et al. 2018 |
| 87 | history of hypertrophic cardiomyopathy, short stature, developmental delay, and distinctive facies | *LZTR1* increased usage of an exon within intron 16<br>zScore: -1.55, FC: 0.85, p value: 1.09e-1 | c.1943-351G>C | Lionel et al. 2018 |
| 91 | Global developmental delay, autism spectrum disorder, epilepsy, atrial septal defect | *PPP2CA* decreased expression<br>zScore: -3.51, FC: 0.88, p value: 6.97e-4 | Not detected | Lionel et al. 2018 |
| 1035 | Constitutional overgrowth, autistic spectrum disorder, ADHD, pectus excavatum, advanced bone age | *NFIX* (Marshall or Sotos AD): increased usage of alternate transcript and NSD1 (Sotos): 9 bp shortening of exon 22 | Not detected | Stavropoulos et al. 2016 |
| 1058 | ZTTK syndrome | *CEP120* decreased expression and increased usage of a splice isoform targeted for nonsense mediated decay<br>zScore: -4.74, FC: 0.76, p value: 4.15e-7 | Not detected | Stavropoulos et al. 2016 |
| 1085 | Epilepsy, autism spectrum disorder, behavioral disorder | *PTK2B* decreased expression and retention of intron 27<br>zScore: 5.97, FC: 1.68, p value: 1.56e-10 | c.523+1G>A | Stavropoulos et al. 2016 |
| 1101 | Profound global developmental delay, spastic quadriplegia, Acute rhabdomyolysis | *B4GALT2* decreased expression<br>zScore: -3.40, FC: 0.45, p value: 8.30e-4 | Not detected | Stavropoulos et al. 2016 |

***Case 18:*** Decreased expression of the *UGDH* gene, which is associated with AR Developmental and Epileptic Encephalopathy 84, was identified as an extreme outlier (z-score – 4.15, fold change 0.62, p value 7.50e-05) in a male with global developmental delay and epileptic encephalopathy (Figure 3A). The UGDH protein is involved in the biosynthesis of extracellular matrix components and loss-of-function variants result in epileptic encephalopathy with variable degrees of developmental delay (51). No variant was detected in the GS data.

**Figure 3.**
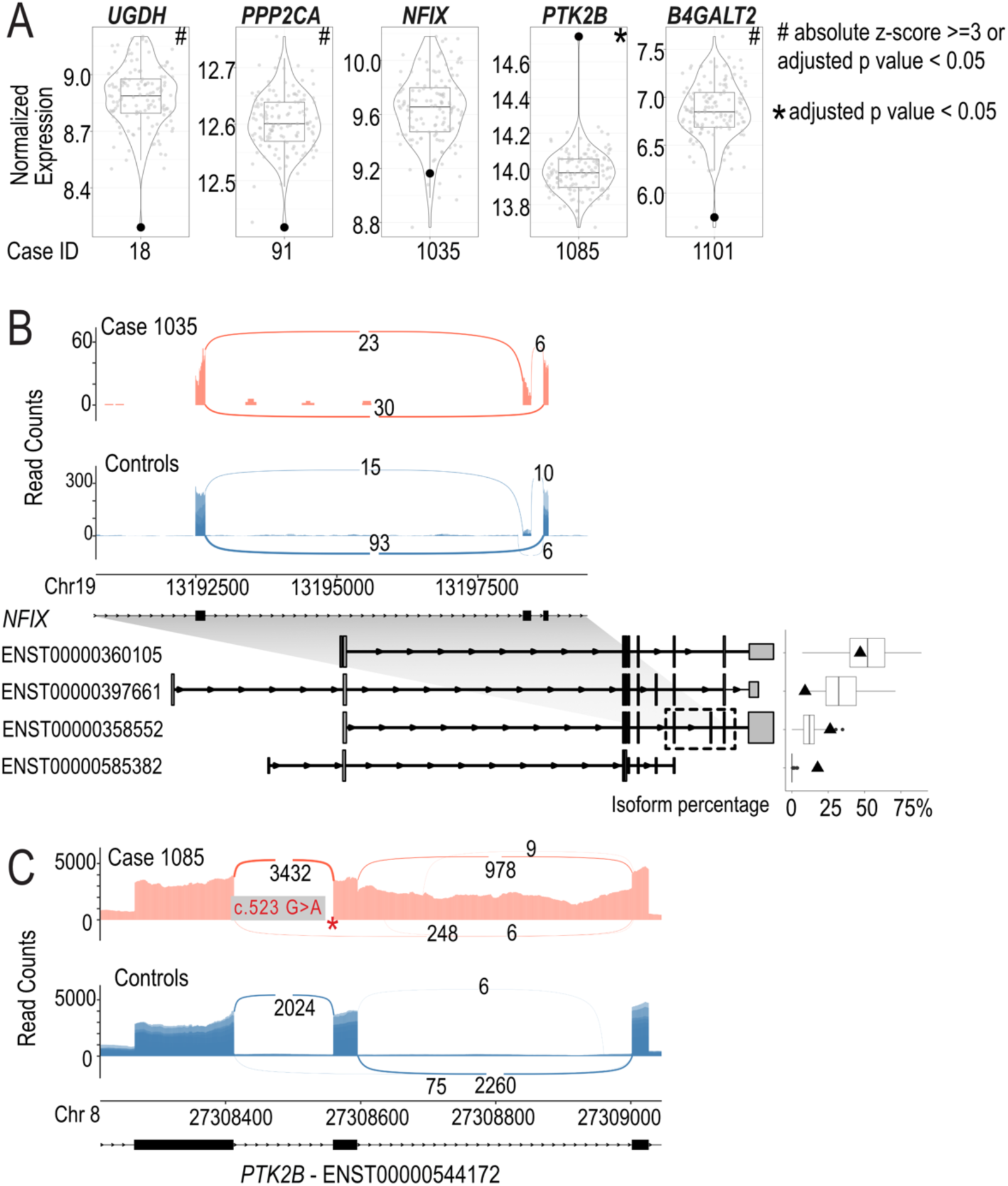
Aberrant splicing events and expression of candidate genes identified using RNA-seq. A. Violin and boxplot showing normalized gene expression of selected candidate genes across the cohort. Y axis: log2 normalized read counts of the target gene in all samples in the cohort. Each dot represents one sample. Highlighted dot represents the case of interest. Case IDs are shown below each plot. Number sign: the gene is considered an outlier in the analysis (absolute z-score >=3 or adjusted p value < 0.05); asterisk: the gene is detected as a significant outlier (adjusted p value < 0.05). **B. Case 1035.** Sashimi plot showing an exon skipping event in *NFIX* in case (red track) and control samples (blue track, n = 10, randomly selected from the cohort). Bottom left: Structures of relevant transcripts. Exons shown in the sashimi plots are highlighted with a dashed box. Bottom right: Boxplot showing the percent isoform usage (“IsoPct” from RSEM output) of the corresponding transcript across the cohort. Each dot represents a sample. Isoform usage of the case is highlighted as a triangle. Only the transcripts detected in the case are shown. **C. Case 1085**. Sashimi plot showing an intron retention event in *PTK2B.* The DNA variant is shown with an asterisk.

***Case 91:*** Decreased expression of the *PPP2CA* gene (Protein Phosphatase 2, Catalytic Subunit, Alpha Isoform associated with AD Houge-Janssens syndrome 3) was identified as an outlier (z-score: -3.51, FC: 0.88, p value: 6.97e-4) in a female with global developmental delay, autism spectrum disorder, epilepsy, atrial septal defects (Figure 3A). No variant was detected in the GS data.

***Case 1035:*** Splicing outlier analysis identified increased usage of exon 9 of the *NFIX* gene (NM_002501) in the blood sample obtained from a male with constitutional overgrowth, autistic spectrum disorder, pectus excavatum, and advanced bone age (Figure 3A and 3B). *NFIX* encodes a transcription factor and is associated with AD Marshall-Smith syndrome and Malan syndrome (Sotos syndrome 2) and is highly expressed in brain cerebellum. No variant was detected in the GS data.

***Case 1085:*** Splicing and expression analysis identified the *PTK2B* gene as a candidate gene due to an intron retention event in intron 27 in a male with features of autism, behavioural issues, obesity, epilepsy and growth hormone deficiency. This intron retention is predicted to result in a frameshift (NM_004103.4) (Figure 3A and 3C). The *PTK2B* gene showed significantly increased expression (z-score 5.97, fold change 1.68, p value 1.56e-10). A re-examination of the GS data revealed a novel variant at +1 splice donor site of intron 27 (c.523+1G>A) that explains the intron retention. The function of the *PTK2B* gene is associated with signaling neuropeptide activated receptors or neurotransmitters and may also provide a mechanism for a variety of short- and long-term calcium-dependent signaling events in the nervous system (55,56). *PTK2B* has been identified as a risk factor for Alzheimer’s disease by multiple studies and the mechanism of some susceptibility alleles has been attributed to splicing alterations (57–59). One Alzheimer’s Disease-risk allele (rs28834970-C) present in this individual is associated with higher protein expression of PTK2B in monocytes (60). In the GTEx whole blood expression dataset rs28834970-C is also associated with moderately increased expression of *PTK2B* (61). This raises the possibility that the individual’s phenotype is the result of two independent molecular events where the common disease-associated SNV (rs28834970-C) upregulates the expression of the aberrant frameshift containing transcript. Since it was not possible to phase these variants, the relevance of this proposed mechanism remains to be seen.

***Case 1101:*** Decreased expression of the *B4GALT2* gene (UDP-GAL: beta-GlcNac beta- 1,4-galactosyltransferase, polypeptide 2) gene was found in blood isolated from a male with profound global developmental delay, spastic quadriplegia and acute rhabdomyolysis (z-score: - 3.40, FC: 0.45, p value: 8.30e-4) (Figure 3A). Although the *B4GALT2* gene shares 55% amino acid identity with the *B4GALT1* gene, which is associated with Congenital disorder of glycosylation, type IId, the role of *B4GALT2* in disease is not known, and further analysis will be required to understand the possible role of the aberrant expression of the gene and the role it may play in relation to the phenotype of this individual. No variant was detected in the GS data. Overall, the application of RNA-seq to cases with no DNA findings resulted in candidate genes related to participant phenotypes in 5 out of the 73 cases. We also identified two cases where RNA-seq provided alternative diagnosis or additional variants of interest:

***Case 87:*** in a male with history of hypertrophic cardiomyopathy, short stature, developmental delay, and distinctive facies in keeping with Noonan syndrome, we detected increased usage of a 117bp exon within intron 16 of the *LZTR1* gene which is predicted to result in an in-frame insertion of 36 amino acids (NM_006767.4; Figure 4A). Within this exon, we also detected an alternative splice acceptor site that is predicted to result in a premature stop site (Figure A). We identified a homozygous rare deep intronic variant c.1943-351G>C that is likely the cause of these two aberrant splicing events. All of the RNA-seq reads aligning to this exon originated from this variant. Although an *EPG5* variant had been detected by GS (Supplementary Table 2), this *LZTR1* finding was a stronger match to the individual’s clinical presentation and was considered to be diagnostic.

**Figure 4.**
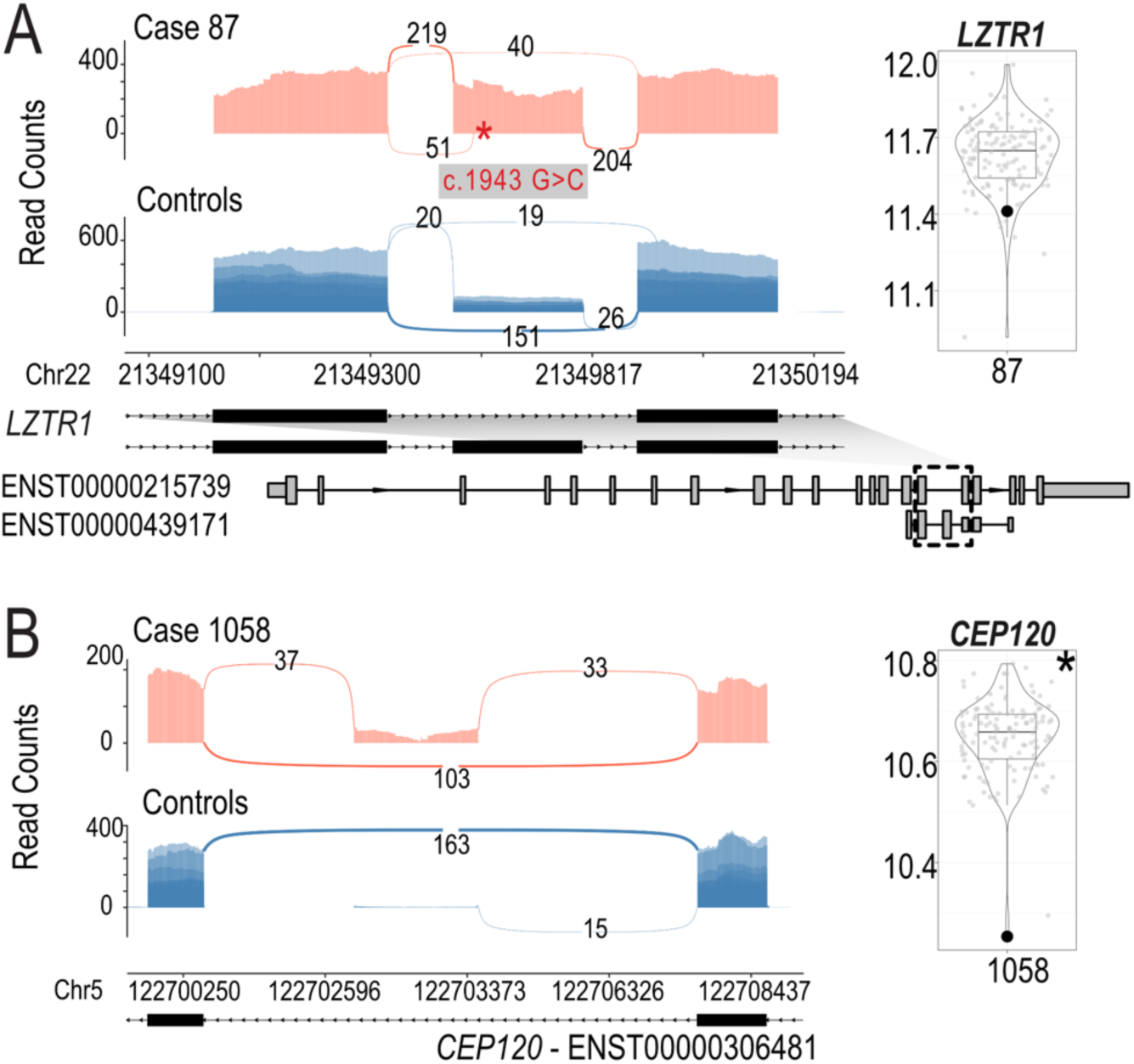
Aberrant splicing events and expression of candidate genes identified using RNA-seq. A. Case 87. Sashimi plot showing the inclusion of a rarely used exon within intron 16 in *LZTR1* and violin plot showing the decreased expression of *LZTR1*. Models of relevant isoforms are shown below. **B**. Case 1058. Sashimi plot showing an exon skipping event in *CEP120* and violin plot showing decreased expression of *CEP120*.

***Case 1058:*** We detected a significant decrease in the expression of the *SON* gene (z-score -3.50, fold change 0.80, p value 6.00e-04) and no effect on splicing in blood obtained from a male who presented with global developmental delay, hypoplastic toes, syndactyly and macrocephaly (Figure 4B). We also detected the *SON* (NM_138927.2) c.3476del (p.Pro1159Arg*9) variant, associated with AD ZTTK syndrome, which was independently identified during DNA re- analysis of the case (18). This demonstrates the ability of an RNA-first approach to detect clinically relevant RNA events even if the DNA variant is not known at the time of analysis. We also identified another candidate gene *CEP120*, which is associated with Joubert syndrome (52), with a novel exon between exon 18 and 19 (Figure 4B) and decreased expression (z-score -4.54, fold change 0.76, p value 4.15e-7). No DNA variants were identified in GS analysis. However, it has been demonstrated that a 57bp deletion overlapping a pair of Tigger4 transposons within intron 17 causes expression of this novel exon (53). We see evidence of this deletion in the RNA-seq data but the zygosity is unclear (Supplementary figure 4). The SON protein regulates the splicing of centriolar protein genes including *CEP131* (54). It is possible that SON also regulates the splicing of *CEP120*, together with the *CEP120* intronic deletion, resulting in the observed reduction in gene expression and potentially contributing to the complexity of the individual’s phenotype.

## DISCUSSION

In this study we benchmarked the diagnostic utility of RNA-seq in a well-studied pediatric rare disease cohort (SickKids Genome Clinic) that was established for assessing the clinical utility of genome sequencing. This unique cohort allowed us to perform a follow-up assessment of blood RNA-seq focussing on two clinical testing scenarios. In the first scenario, RNA-seq was used to provide functional data for variants that had been identified by GS and were considered diagnostic for the participant’s phenotype. Using RNA expression as functional readout of genetic changes is a rationale and practical way to help confirm diagnoses and suggest pathogenic mechanisms. Assigning a molecular mechanism to a genetic disease diagnosis is a requisite for bespoke genetic therapy development (62). In the second scenario, we aimed to identify candidate genes using a transcriptome-first analysis. Our work showcases a variety of cases where RNA-seq effectively facilitates the diagnosis and will contribute to the establishment of a clinical guideline. When comparing the RNA-seq data to *in silico* prediction algorithms, the results are concordant for the majority of the SNV variants analyzed. However, there were five notable exceptions. The homozygous c.2600_2601delTA p.(Leu867*) [NM_017733.3] variant in the *PIGG* (Case 64) gene, which was predicted to result in NMD. We had expected that a homozygous nonsense variant would have resulted in a pronounced decrease of expression, but we only observed moderately decreased levels (z-score -2.34, fold change 0.88, p value 0.028, which it did not pass our cutoff filter). This illustrates the impact that technical gene expression cut-offs and filtering algorithms will have on potentially biologically complex, yet still meaningful observations. A combined approach of using lenient cutoffs for well-studied genes and more stringent cutoffs for novel disease genes could help alleviate this issue. It also reinforces the importance of RNA-seq for functional categorization of variants from a therapy development prospective, as patients with predicted NMD associated SNVs are considered for NMD read through therapy, which would be appropriate or effective if RNA analysis shows a lack of predicted transcript degradation.

Similarly, two missense variants, (Case 1029*: PIK3R2* c.1117G>A (p.Gly373Arg) [NM_005027.4]) and (Case 1093*: COG5* c.1205C>T (p.Ser402Leu) [NM_006348.3]) each 7bp and 3bp respectively from the splice acceptor sites were computationally predicted to affect splicing. However, RNA-seq data showed no effect on splice site usage. Lastly, frameshift variants in the *PANK2* (Case 1016) and *SMARCB1* (Case 1008) genes, were predicted to result in NMD but we showed a splicing impact. Our analyses show that *in silico* approaches can fail to predict the specific splicing outcome, and that at least 14% of variant analyses would benefit from RNA-seq data. RNA-seq data provides individual-specific, empirical information on specific splicing mechanisms (exon skip, intron retention) which can be useful even when canonical splice site variants are encountered. This observation echoes the conclusions from other published work (63) and highlights the need for obtaining more pediatric RNA-seq data to improve *in silico* RNA splicing prediction algorithms.

Our study also highlights the value of using RNA-seq in cases with large CNVs identified by CMA. Consistent with findings by others, we showed that gene expression changes cannot be accurately predicted from gene copy number (identified by array or GS) alone (41,42,44,46,47). The examples that we present showed that RNA-seq provides information on dosage compensation at the transcriptional level for individual genes within CNVs and complex chromosomal re-arrangements. For example, decreased dosage of a combination of genes in individuals with 22q11 deletion syndrome is attributed to the complex and variable 22q11 deletion syndrome phenotypes (64,65). By measuring the expression change of every gene in an unbiased manner, RNA-seq could provide further insights into the pathogenicity of CNVs and pinpoint sets of genes contributing to inter-individual differences in phenotype. These data will be relevant for the more complex phenotypes and add a nuanced interpretation to genome sequencing findings. However, additional information on gene expression and epigenomic features in disease-relevant and developmental stage specific samples will be needed to understand the pathogenicity of sets of genes affected by CNVs (41,47,66).

Using an RNA-centric approach on the second cohort of individuals who did not receive a genetic diagnosis by GS or CMA, we identified aberrant splicing or expression events related to the primary clinical indication in ∼10% (7/73) of the cases. However, it remained challenging to identify a causal DNA variant leading to these clinically relevant RNA phenotypes, especially for gene expression outliers. The yield seen in this RNA-centric approach may be the result of DNA variants that are poorly annotated, for example regulatory, intronic or intragenic areas of the genome or structural changes not identified by short read sequencing. In addition, both the genomic and transcriptomic analyses only included genes with known disease or HPO associations and would not have been able to identify molecular changes in novel disease genes. We identified a diagnostic variant in *LZTR1* (case 87) before a DNA variant had been found, underscoring the ability of RNA-seq to identify splicing and expression outliers and the causative DNA variant.

Our study highlights some of the challenges facing the routine use RNA-seq in a clinical genetic setting identified in previous studies (4–15). Firstly, using blood as a proxy tissue for non-blood related clinical indications may fail to detect gene expression levels and cell-type specific splicing. Even in the cases where robust gene expression was detected in blood, different epigenetic processes in the disease-relevant cell type could change the outlier gene expression and splicing results which in turn could change the clinical interpretation. More sensitive targeted technologies such as targeted PCR-based approaches (e.g. (7)) as well as trans- differentiation of accessible cell types from blood or skin into more disease-relevant tissue types (e.g. (67)) will help to overcome this challenge. While we have used our own cohort as a pediatric control population to minimize technical and biological differences, subtle variations may have escaped the identification of outliers under a fixed cutoff. It has been shown that the number of controls have a strong impact on gene expression outlier analysis (11). It is thus critical to collect age-, sex-, and tissue-matched control cohort that are processed using a uniform experimental and computational pipeline. Building and sharing databases of high-quality pediatric RNA expression data is a realistic solution to achieve this.

In conclusion, our data show the utility of peripheral blood RNA-seq in assessing the genetic underpinnings of a diverse range of disorders. By utilizing a clinical-grade, automated version of a standard RNA-seq library preparation methodology on whole blood samples we show that RNA-seq is sufficiently sensitive and reproducible to provide functional evidence for variants identified by genome sequencing. In this context, our results support using RNA-seq diagnostics in parallel with genomic sequencing. Such benefits could include increasing the confidence in, and reducing the time spent reporting GS findings. Furthermore, providing critical clarification of variant impact, a critical step for assessing and designing individualized genetic medicines (62). However, from a health economics perspective, it is unclear if a dual DNA/RNA testing regime is financially justified and how it compares to reserving RNA-seq for individuals for whom GS did not result in a conclusive finding. Nevertheless, expediating the production of a comprehensive and diverse reference set of pediatric blood gene expression profiles will enhance RNA-based genetic diagnoses and enable new algorithms for precision child health.

## Acknowledgements

We are grateful for the individuals and their families who participated in this study. Special thanks to Sergio Pereira and Karen Ho at The Centre for Applied Genomics, Lisa Knapp at Agilent and Howard Cukier at New England BioLabs for their support in the development and optimization of the automated RNA-seq platform, and all past contributors to the SickKids Centre for Genetic Medicine and Genome Clinic project. Funding was provided by Genome Canada (OGI-158 to MB, AS, JJD, MDW). MBrudno and DJS have equity in PhenoTips. SWS is supported by a Northbridge Chair in Paediatric Research and JJD is supported by the Mogford Campbell Family Chair of Paediatric Neurosciences.

## Contributions

AS, JJD, LK, MDW, MBrudno, and MSM obtained funding, conceived the study design and supervised the work. HH, AKR, AC, MBrudno, AS, MDW were responsible for data analysis pipelines. LK, KEY, HH, GC, SBarnes, AS, DJS, MSM, CRM were involved in data interpretation and clinical assessments. HH, AKR, AC were responsible for RNA-seq quality control analyses. MBraga and MGB generated the RNA-seq libraries as well as testing and optimizing the automated RNA-seq library preparation method. SWS coordinated and KEY led the implementation of the automated RNA-seq library preparation method at the Centre for Applied Genomics. RDC, SBowdin, RML, SWS, CRM, MSM, DJS represent the SickKids Genome Clinic who obtained, organized and analysed the genome sequencing data. HH, KEY, LK and MDW wrote the manuscript. All authors read and reviewed the manuscript.

## Data availability

The data supporting the current study was consented for research but not for sharing in public databases. Based on the results of our study this dataset will serve as pediatric blood expression control samples for rare disease research. Please contact the corresponding authors if you wish access to this data.

