## Supplementary Figures for "Assessing the diagnostic impact of blood transcriptome profiling in a pediatric cohort previously assessed by genome sequencing"

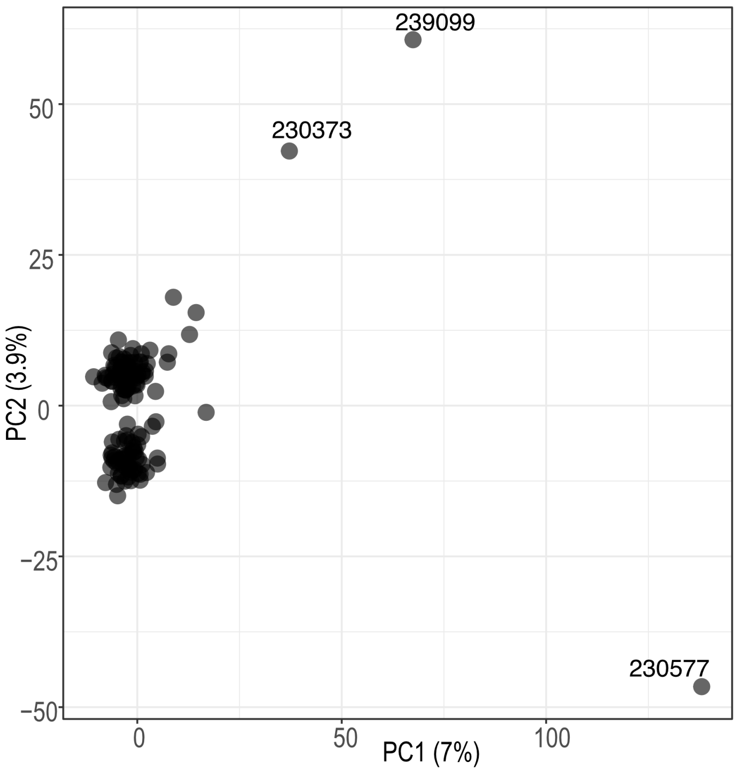


**Supplementary Figure 1. PCA plot showing outlier samples after OUTRIDER normalization.** Each dot represents one sample. The IDs of the three outlier samples are shown. Log2 transformed “normcounts” from OUTRIDER output was used for the PCA analysis.

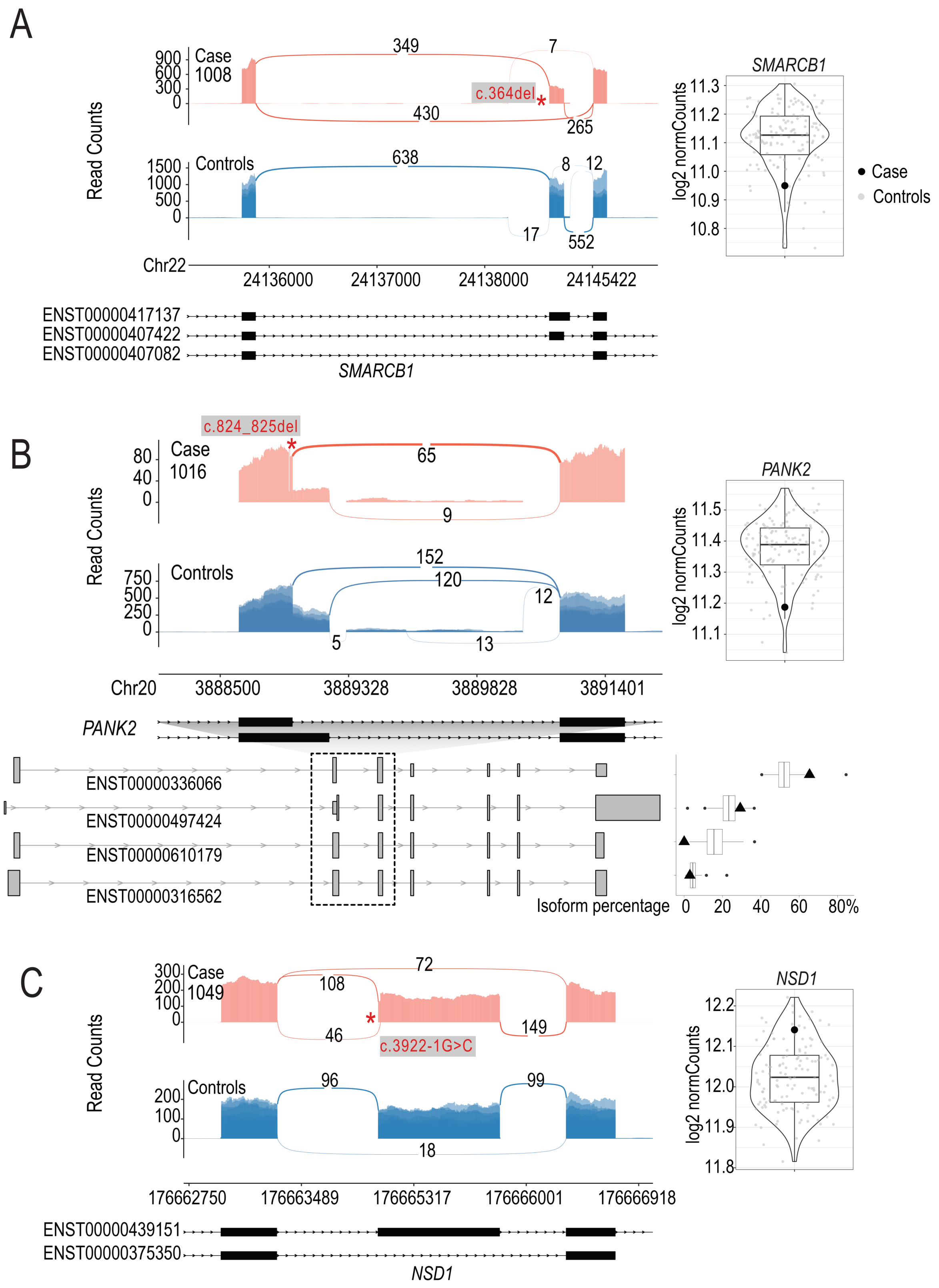


**Supplementary Figure 2. Transcriptional impact of candidate variants identified with WGS/WES. A. Sashimi plot showing an exon skipping event in gene *SMARCB1.*** Red:

proband; blue: control samples (n = 10, randomly selected). Y axis: read counts. Violin plot showing non-significant decreased expression of *SMARCB1*. Y axis: log2 normalized read counts. Black dot: cases in which the corresponding gene was a candidate; Grey dot: rest of the cohort as control samples. The DNA variant is shown with an asterisk **B**. **Sashimi plot showing increased usage of a shorter exon in gene *PANK2* and the pathogenic variant (two base pair deletion).** Violin plot showing non-significant decreased expression of *PANK2*. Bottom left: Structures of relevant transcripts. Exons shown in the sashimi plots are highlighted with a dashed box. Bottom right: Boxplot showing the percent isoform usage ("IsoPct" from RSEM output) of the corresponding transcript across the cohort. Each dot represents a sample. Isoform usage of the case is highlighted as a triangle. **C.** S**ashimi plot showing increased usage of an isoform that skips impacted exon and 5bp shift of the impacted splice acceptor site in gene *NSD1*.** Violin plot showing no notable impact on *NSD1* expression.

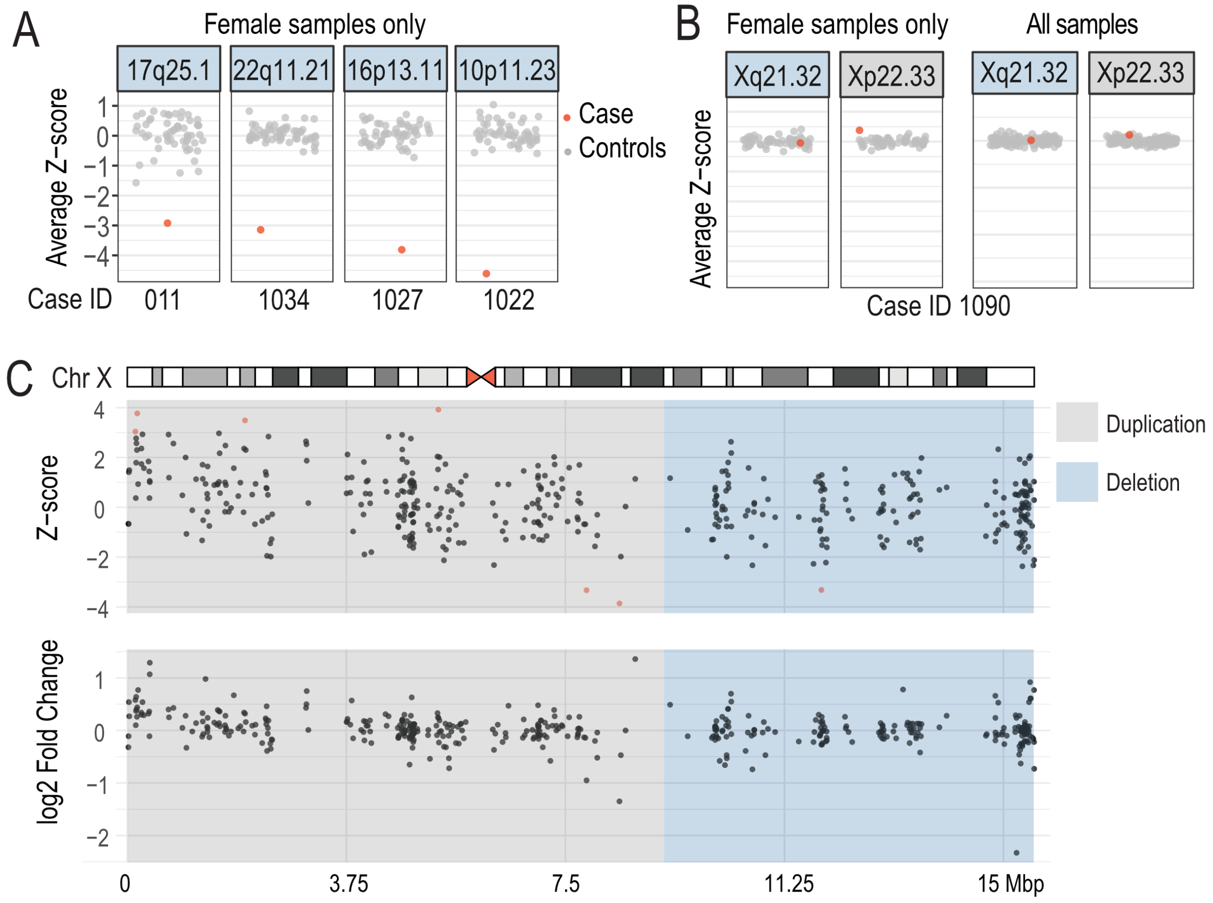


**Supplementary Figure 3. Modulation of gene expression by CNVs analyzed using female samples only.** Z-score summary of genes within **A.** pathogenic autosomal CNVs and **B.** chromosome X CNVs. Analysis performed with female samples only or all samples. Y axis: average z-score of genes in pathogenic CNVs. Each dot represents one sample. Red: sample in which the pathogenic CNV is detected. Panel header: Sample ID and CNV coordinate. C**. Z-scores and fold-changes of all genes on chromosome X in case 1090.** An Ideogram of human chromosome X is shown. The heterozygous duplicated region is highlighted in grey and deleted region in grey. Gene start is used as the proxy for gene location. Genes with an absolute z-score >= 3 are highlighted in red.

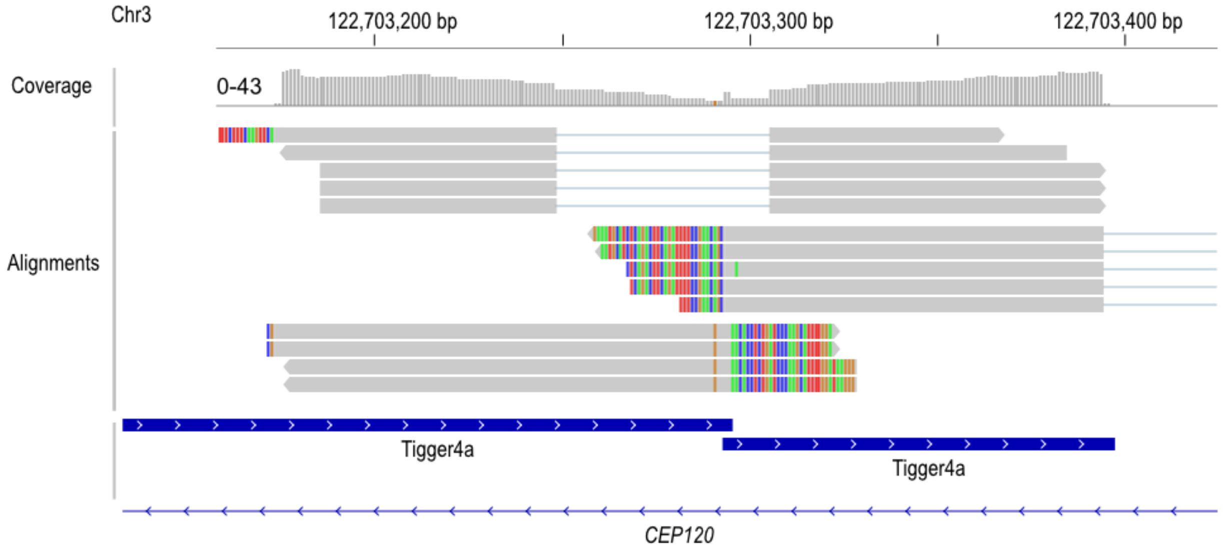


**Supplementary Figure 4. IGV screenshot showing read alignment and coverage across the novel exon in *CEP120*.** From top to bottom: 1) genome scale; 2) coverage track with the scale bar shown on the side; 3) Example aligned reads showing the 57 bp deletion (first group) and misalignment due to repetitive elements (second and third groups). Colored bases represent soft-clipped bases. 4) transposable elements and gene model track.
